# Validation of a Novel Algorithm for Automated Detection and Quantification of Choroidal and Retinal Pulsation on Video Indocyanine Green Angiography

**DOI:** 10.64898/2026.08.10.26360105

**Authors:** Niroj Kumar Sahoo, Utkarsh Doshi, Giulia Gregori, Diana Flores-Peña, Marco Lupidi, Kiran Kumar Vupparaboina, Jay Chhablani

## Abstract

**Purpose:** To validate an automated pipeline to detect and quantify focal retinal and choroidal pulsation areas that are synchronous with the cardiac cycle in video indocyanine green angiography (ICGA).

**Design:** Retrospective, observational, hypothesis-generating validation study Subjects, Participants: Consecutive patients with a diagnosis of central serous chorioretinopathy (CSCR) in one or both eyes.

**Methods:** Videos were acquired on Heidelberg HRA+OCT. The pipeline consisted of three steps: signal extraction, foci detection, and quantification. After registration of the constituent frames, each pixel’s intensity signal was analyzed at the presumed cardiac frequency (tested from a sample of three detectable frequencies). A synchrony score combining local phase coherence with oscillation amplitude was then derived and computed using a standard deviation (σ) above each video’s background oscillation value. Two masked graders marked the retinal and choroidal pulsation areas twice. We compared detection of the pulsation areas against grader consensus using a receiver operating characteristic curve (using multiple grid sizes to divide the scan area) and, separately, using a signal-based area-reduction method to obtain an optimum σ value.

**Main Outcome Measures:** Agreement between the automated algorithm and human graders in detection of pulsation foci, and the optimum threshold multiplier (σ).

**Results:** We studied 20 ICGA videos from 20 eyes. At the 16-pixel grid size, the pipeline achieved a mean area under the curve (AUC) of 0.914, sensitivity of 0.86, and specificity of 0.80. Grader agreement improved with larger grid size, reaching substantial-to-strong levels for choroidal annotations. The two independent validation methods demonstrated similar σ values that differed by 0.62σ, supporting σ=4.0 as the optimum value.

**Conclusions:** We report the first automated method to quantify retinal and choroidal vascular pulsation on video ICGA. It measures pixels that oscillate over time with the presumed cardiac cycle and works reliably at the spatial scale (grid level) where experts agree. Pulsatile hemodynamics may add a new vascular biomarker for glaucoma, diabetes, hypertension, and pachychoroid diseases.

## INTRODUCTION

The uniqueness of the eye as an organ lies in the fact that the vasculature can be directly and non-invasively visualized. This makes retinal and choroidal vasculature invaluable not just for diagnosing ocular disease but also for understanding systemic health. While retinal vascular changes like retinal arterial narrowing, arteriovenous nicking, and microaneurysms have been associated with hypertension, diabetes, and cardiovascular mortality for decades,1, 2 the choroid provides metabolic support to the outer retina and accounts for approximately 90% of total ocular blood flow.3, 4

The very richness of the ocular vasculature presents its own analytical challenge.5 Clinically useful biomarkers have been extracted from each of the vasculature using several generations of imaging technology, including fundus fluorescence angiography (FFA), indocyanine green angiography (ICGA), optical coherence tomography (OCT), and OCT angiography.6 However, the current armamentarium does not adequately capture the dynamic behavior of the vasculature, specifically the oscillation of individual vessel segments. In systemic circulation, arterial pulsatility has long been used as a clinical biomarker to assess stiffness and vascular resistance across multiple organs.7, 8 In the eye, however, pulsatile vascular behavior has been unexplored as a quantitative biomarker, primarily due to two intrinsic challenges. First, the signal is subtle, providing brightness oscillations in angiograms that are difficult to distinguish from background noise, dye front movement, and imaging artifacts. Second, the spatial complexity of the choroidal vasculature, with overlapping vessel layers, makes it extremely challenging for a human observer to reliably identify and localize individual pulsating foci. As a result, choroidal pulsation foci have been noted anecdotally in ICGA studies but have not been appropriately quantified. The clinical relevance of these pulsatile signals in the choroid has been described in recent work by Cheung et al., who combined video ICGA with sequential frame subtraction in patients with CSCR and pachychoroid disease.9 The authors demonstrated that filling of large choroidal veins, particularly in watershed zones and at intervortex venous anastomoses, occurred in a pulsatile fashion. However, this technique involves manual processing, is limited to the dye-bolus phase of the angiogram, requires a complete absence/presence of dye to enable detection (smaller fluctuations in dye intensity may be missed), and does not provide quantitative metrics. The present study addresses this gap by describing and validating a fully automated computational pipeline for detection and quantification of focal retinal and choroidal pulsation in video ICGA recordings. The pipeline operates by computing for every pixel in the registered angiographic video and analyzing whether pulsation is coherent with a presumed cardiac cycle and is of measurable magnitude.

## METHODS

### Study Design Overview

This was a retrospective, observational, hypothesis-generating, and validation study conducted in eyes with chronic CSCR and available ICGA videos, and with proven visible retinal and choroidal pulsation. The study was conducted in accordance with the Declaration of Helsinki and approved by the institutional ethics committee. Consent was waived due to the retrospective study design. The pipeline consisted of three basic steps: signal extraction, focus detection, and quantification.

### Dataset

A sample of 20 videos from 20 eyes with at least one retinal focus and one choroidal focus of pulsation, as detected by the graders (GG and DFP), was selected for analysis. All videos were acquired on HRA + OCT (Heidelberg Engineering, Heidelberg, Germany, using a 55° field of view, and were registered using the built-in function in the HEYEX system to reduce motion artifacts. Early-phase angiograms of approximately 10-15 seconds at 9 frames per second were selected. The various stages of the analysis have been listed below:

### Signal extraction

The videos were first divided into individual frames, and dark frames (originating from blinks and gross motion) were discarded. A motion-stable fluorescence window (excluding the early unstable transient dye-flush phase) was used for all analysis. Enhanced Correlation Coefficient (ECC) alignment was performed for registration and was skipped when estimated displacement was <2 pixels. Edges of the video frames with dark pixels were excluded automatically. A manual mask was also painted over peripheral zones where edge motion could be misread as oscillation.

Within the stable window, each pixel’s mean-subtracted, band-limited (0.5–3.0 Hz) intensity signal was projected onto a trial frequency f via a windowed complex demodulation, yielding amplitude and phase values. The most likely cardiac frequency f was selected from a range of f ranging from 0.7 (42 beats per minute) to 2.50 (150 beats per minute) Hz. At each frequency, the phase-locking value (PLV) was calculated, which measures how well a group of pixels agree on that timing. If the groups have similar frequency, the PLV is close to 1, and if their timings are random, the PLV is close to 0. The group PLVs were visualized only for the pixels exceeding the 70th percentile amplitude. Up to three frequencies were reviewed against each video’s pixel distribution by a grader (NKS), and the physiologically plausible f [determined by comparing the phase locking value, the biological plausibility of the specific frequency as a cardiac beat, and the degree of edge motion detected at that frequency (panels with high values coming from the edges of the video are more likely to be false)] was retained for all downstream analysis of that video (Figure 1). At the selected frequency, we derived a local phase-coherence map and a rescaled amplitude map, combined into a weighted synchrony score (Sp), which is low unless a pixel is both locally phase-consistent and shows measurable oscillation amplitude (Figure 2C, D).

**Figure 1:**
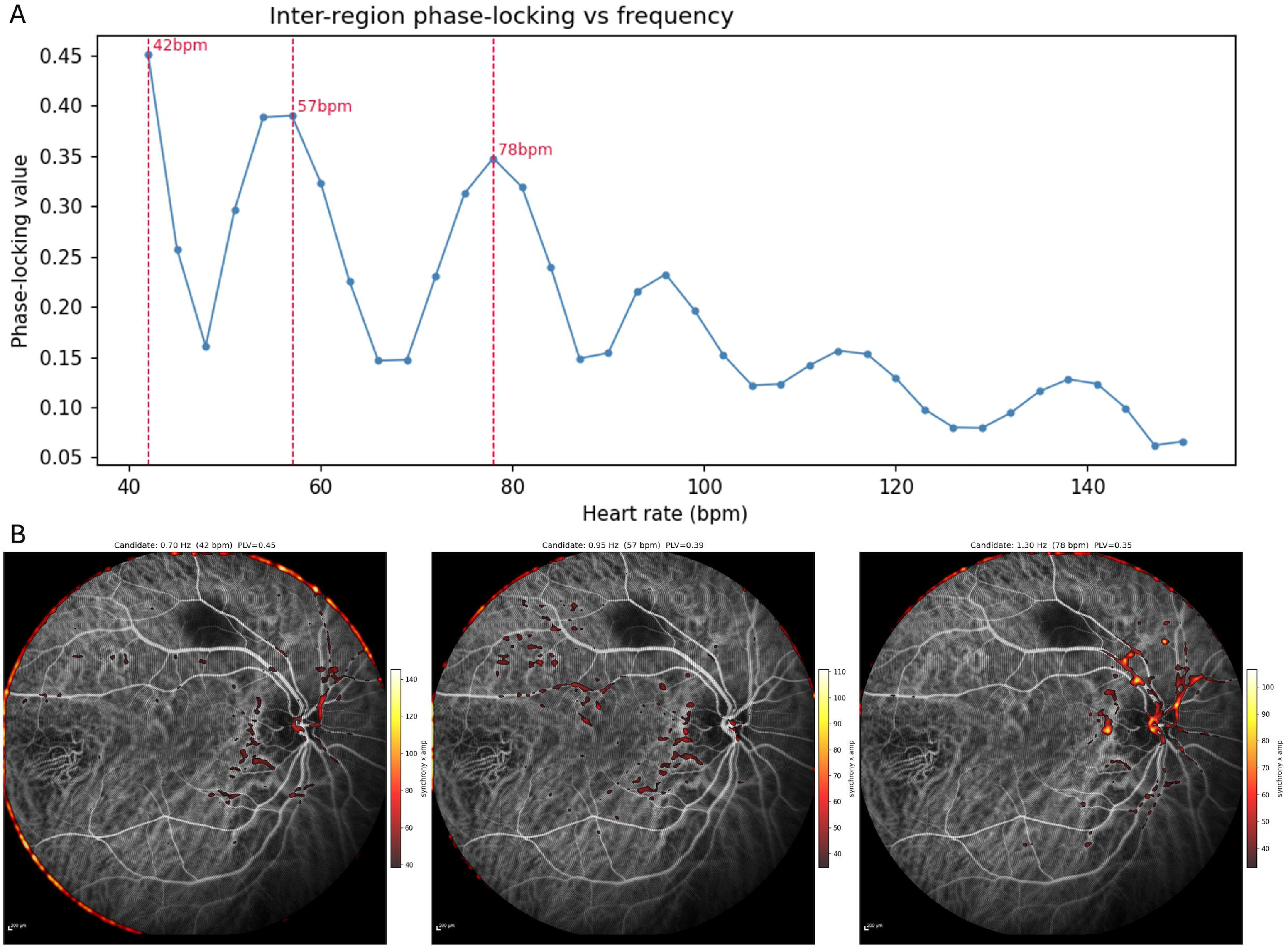
(Processed using Supplementary Video 1) Cardiac-frequency candidate selection for a representative eye. (A) Phase-locking value (PLV) as a function of heart rate, computed across the highest-amplitude non-excluded pixels over the swept frequency range (54–150 bpm). Dashed vertical lines mark the local PLV maxima identified as candidate cardiac frequencies. (B) Synchrony × amplitude heatmaps (Gaussian-smoothed for display) at each candidate frequency marked in (A), ordered left to right by increasing frequency; panel titles report each candidate’s frequency (Hz/bpm) and corresponding PLV value from (A). The operator selected the frequency 1.30 Hz (78 bpm) in this case, as it is biologically plausible, has reasonable PLV values, and the panel signals overlay the retinal and choroidal vessels, with minimal signals from peripheral edge motion.

**Figure 2.**
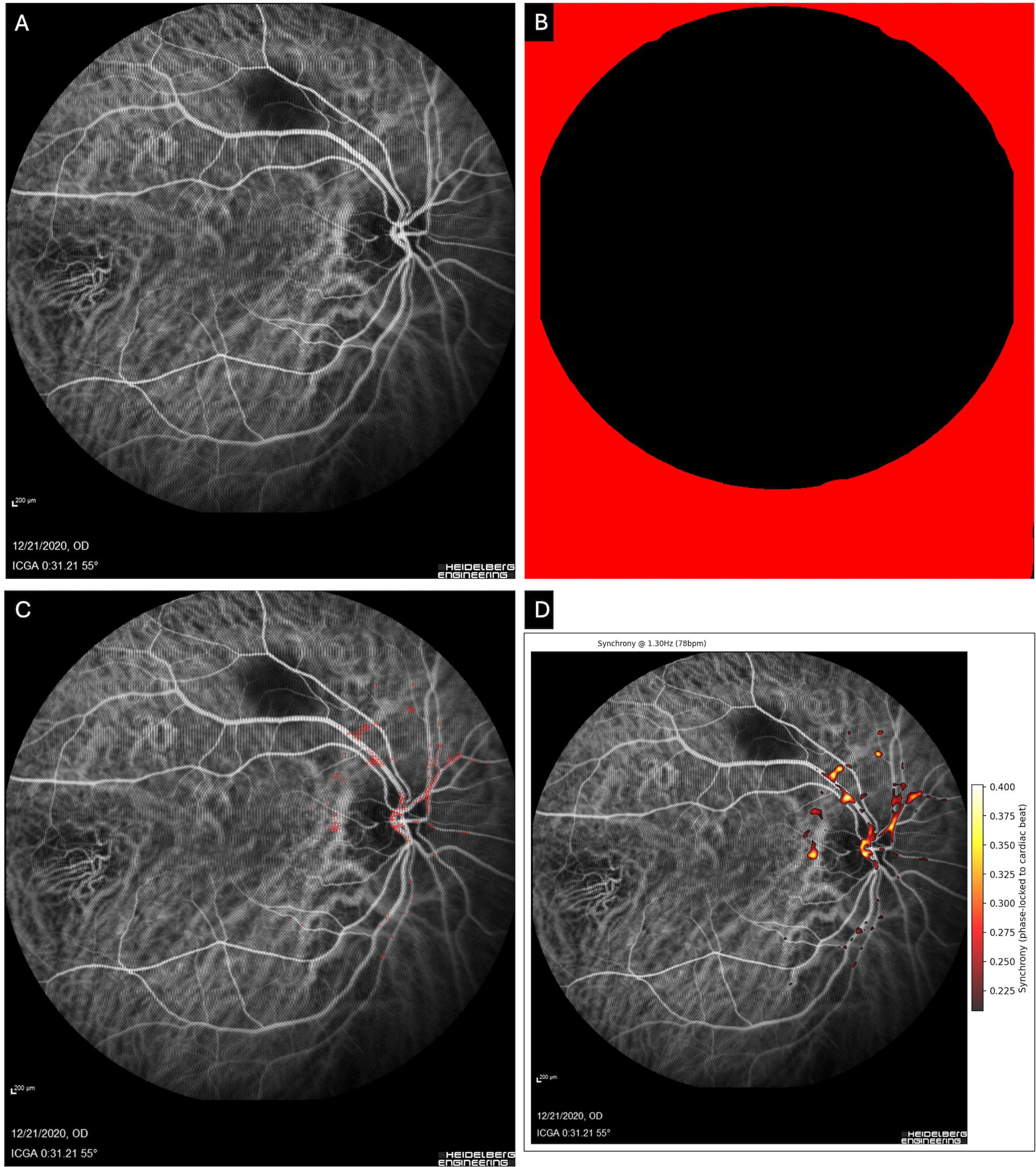
Representative outputs for Supplementary Video 1. (A) Reference frame (last plateau frame). (B) Exclusion mask (red = excluded pixels). (C) Synchrony map (Sp) rendered at threshold σ = 4 that was used for analysis. (D) Smoothened Gaussian filter (Gaussian σ = 3 px) synchrony map for visualization (pixels below threshold of 0.6 masked for clarity).

### Focus detection

A pixel was counted as pulsatile only if its synchrony score Sp rose clearly above the video’s own background. We set the cut-off at θ(σ) = μ + σs, where μ and s are the average and spread of Sp across all valid pixels, and σ sets how far above average a pixel must reach. To keep a few extreme pixels from distorting μ and s, the highest values were capped at the 99th percentile before these were calculated. The parameter σ therefore determines how far above a video’s own background noise level a pixel’s signal must rise before being classified as a detected pulsation and is the sole free parameter governing detection sensitivity in the pipeline. Initially, each active pixel was expanded outward by 8 steps in all directions. If two distinct pixel patches grew close enough to merge, they were combined into one group. After grouping, the gap-filling step was discarded, leaving only the original active pixels for counting. A group was considered a valid focus only if it contained at least 8 of these original pixels; smaller groups were regarded as noise and ignored. Each confirmed focus was then quantified as described in the following section.

### Quantification

Pulsatile extent was expressed as the fraction of valid pixels exceeding threshold and, via an automatically detected 200 µm scale bar, converted to mm² where detection succeeded. Peak/mean amplitudes were reported per focus as strength descriptors.

### Determining the ideal sigma value (σ)

The choice of σ directly determines the sensitivity and specificity of foci detection. A lower σ value lowers the detection threshold relative to the background noise distribution, increasing the likelihood that random fluctuations are misclassified as pulsatile foci (false positives). On the other hand, a higher σ value restricts detection to only the strongest signals, increasing the likelihood that lower-amplitude pulsation is missed (false negatives) (Figure 3). The algorithm was initially set to a default operating threshold (σ=4.0), based on preliminary observations indicating that this threshold most likely represented the most clinically visible pulsation. Thus, further analysis was performed to validate this threshold.

**Figure 3:**
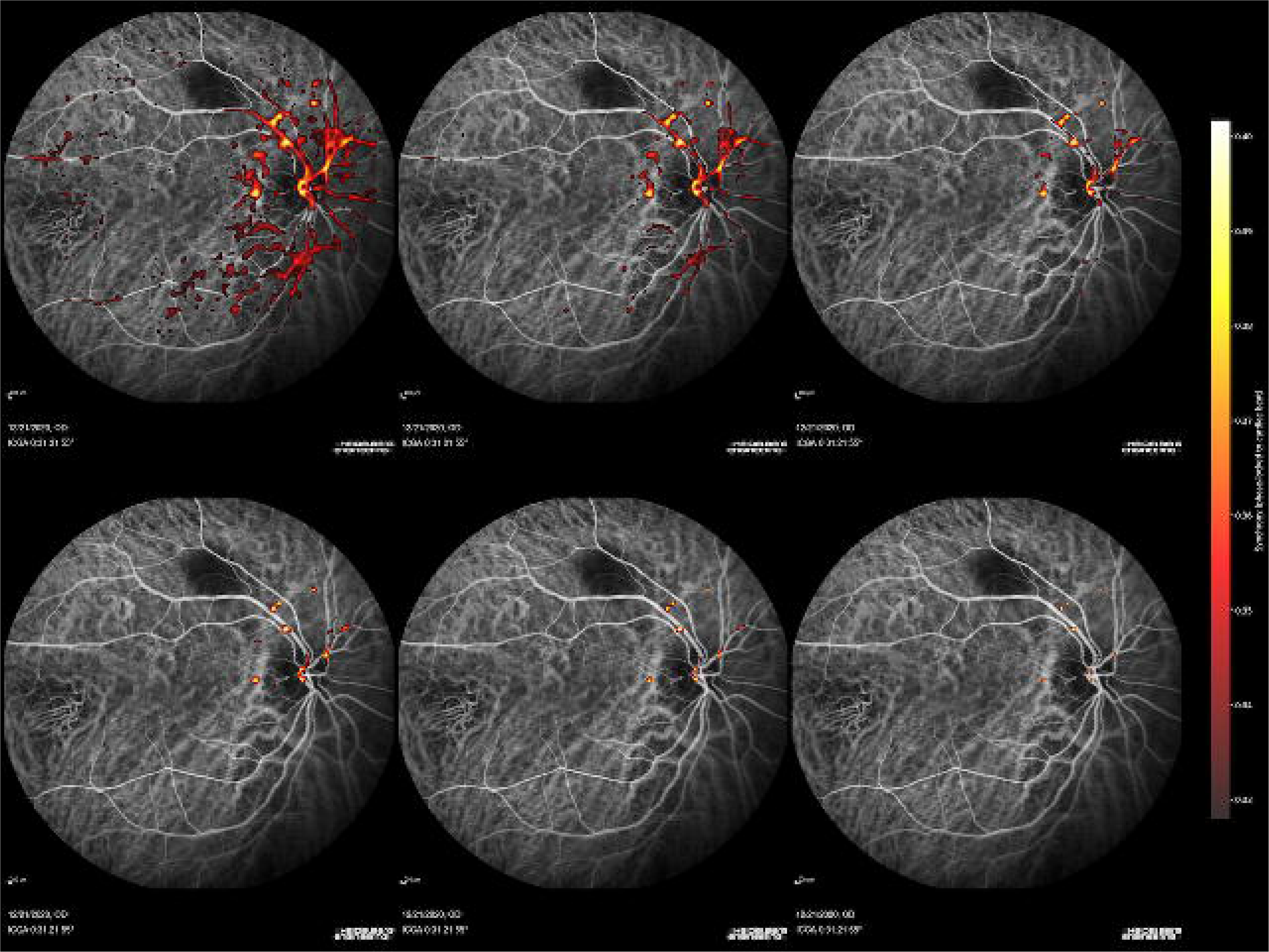
Visual representation (Processed using Supplementary Video 1) (Gaussian-smoothed for display) of the effect of sigma threshold values (σ = 2.0, 3.0, 4.0, 5.0, 6.0, 7.0) on candidate pulsatile area for a representative eye. Panels are arranged in increasing order of σ, left to right and top to bottom. The colored area contracts progressively as σ increases, reflecting a more conservative threshold.

### Validation and statistical analysis

Two graders (GG and DFP) analyzed the videos and manually marked the areas of retinal and choroidal pulsation twice for each video. The videos were randomly assigned, with each grader unaware of their previous markings and those of the other grader. Inter- and intra-rater reliability were assessed via Cohen’s κ and Bland–Altman limits of agreement, separately for retinal and choroidal layers and for inter-rater and pass-to-pass comparisons. Ground truth was defined as the majority (≥2-of-4-pass) consensus of two graders, obtained twice, with 1 week between annotations. Due to possible human-level inaccuracies in pixel-level markings, we adopted a grid-wise framework, comparing box-level positivity (non-overlapping boxes of 8, 16, 32, 64, 128, 256-pixel sizes) between the consensus mask and tested across 27 σ values (1.5 to 8.0, in steps of 0.25) for each box configuration. Sensitivity, specificity, and AUC (trapezoidal rule) per video and box size, with the Youden-optimal σ, were analyzed per combination.

As an entirely independent, annotation-free validation method, we tracked detected pixel areas across the σ values and identified the threshold at which 90% of the removable background area had been eliminated. This threshold was chosen at approximately 90% reduction because at this level, most background noise would have been eliminated, leading to a slower reduction in detection and indicating stable detection. Agreement between this signal-derived operating point and the grid-wise result was taken as a sensitivity analysis. All analyses were implemented in Python 3.14 (NumPy, SciPy, OpenCV, pandas).

## RESULTS

Twenty ICGA videos were processed. The operator-selected cardiac frequency ranged from 78–96 bpm (mean 84 ± 6 bpm). Primary consensus ground truth (≥2/4 passes) was extracted for all 20 videos; mean annotated area was 2,237 ± 756 pixels per video (range 1,202–4,063 pixels).

### Grader Agreement

Grader agreement across all six grid sizes is shown in Table 1 and Figure 4. At the smallest grid size (8px), inter-grader κ was 0.40 ± 0.20 (retinal) and 0.46 ± 0.12 (choroidal), in the fair-to-moderate range. Agreement improved progressively and consistently with increasing box size for both retinal and choroidal annotations. At the 32px size, inter-grader κ was 0.56

**Figure 4:**
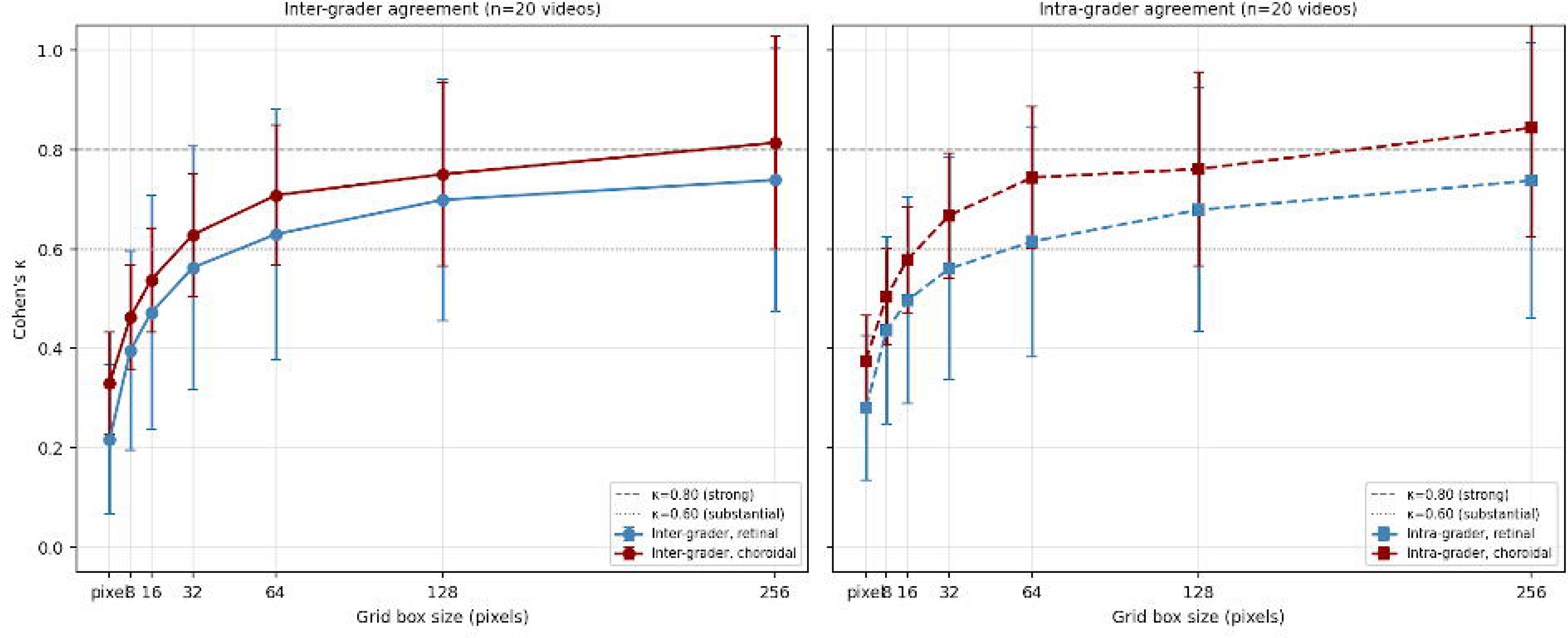
Cohen’s κ vs grid box size, separately for retinal and choroidal annotations (mean ± SD, n=20 videos). Dashed lines at κ=0.60 (substantial) and κ=0.80 (strong).

**Table 1.** Cohen’s κ at all six grid sizes, for inter-grader and intra-grader comparisons, separately for retinal and choroidal annotations.

| Grid size | Inter-grader Retinal $\kappa$ (mean±SD) | Inter-grader Choroidal $\kappa$ (mean±SD) | Intra-grader Retinal $\kappa$ (mean±SD) | Intra-grader Choroidal $\kappa$ (mean±SD) |
| --- | --- | --- | --- | --- |
| 8px | $0.40 \pm 0.20$ | $0.46 \pm 0.12$ | $0.44 \pm 0.20$ | $0.50 \pm 0.14$ |
| 16px | $0.47 \pm 0.24$ | $0.54 \pm 0.13$ | $0.50 \pm 0.21$ | $0.58 \pm 0.14$ |
| 32px | $0.56 \pm 0.25$ | $0.63 \pm 0.14$ | $0.56 \pm 0.23$ | $0.67 \pm 0.15$ |
| 64px | $0.63 \pm 0.26$ | $0.71 \pm 0.16$ | $0.62 \pm 0.24$ | $0.74 \pm 0.16$ |
| 128px | $0.70 \pm 0.25$ | $0.75 \pm 0.20$ | $0.68 \pm 0.25$ | $0.76 \pm 0.21$ |
| 256px | $0.74 \pm 0.28$ | $0.81 \pm 0.22$ | $0.74 \pm 0.28$ | $0.84 \pm 0.23$ |

Values are mean ± SD across 20 videos and all grader pairs. $\kappa$ : $<0.20$ slight; $0.21$ – $0.40$ fair; $0.41$ – $0.60$ moderate; $0.61$ – $0.80$ substantial; $>0.80$ strong.

± 0.25 (retinal) and 0.63 ± 0.14 (choroidal); intra-grader κ was 0.56 ± 0.23 and 0.67 ± 0.15, respectively. At 256px, inter-grader κ reached 0.74 (retinal) and 0.81 (choroidal). Choroidal agreement was consistently higher than retinal at all grid sizes for both comparison types.

Intra-grader κ was slightly higher than inter-grader κ at most grid sizes, indicating that graders were moderately more self-consistent than they were consistent with each other. This progressive improvement with box size demonstrates that both graders reliably agreed on which regions contained pulsation, even when drawn boundaries differed at finer scales.

### Grid-Level ROC

Grid-wise AUC and optimal σ (median across 20 videos) are shown in Table 2 and Figure 5. The 16px grid size achieved the highest mean AUC, closely followed by 32px; both AUC and its consistency (SD) degraded at coarser grid sizes (≥64px). Optimal σ increased with grid size, from 3.25 (8px) to 5.75 (256px). A representative image from a video comparing the combined grading annotations with pixel detection and the optimal σ for different grid sizes is shown in Figure 6.

**Figure 5:**
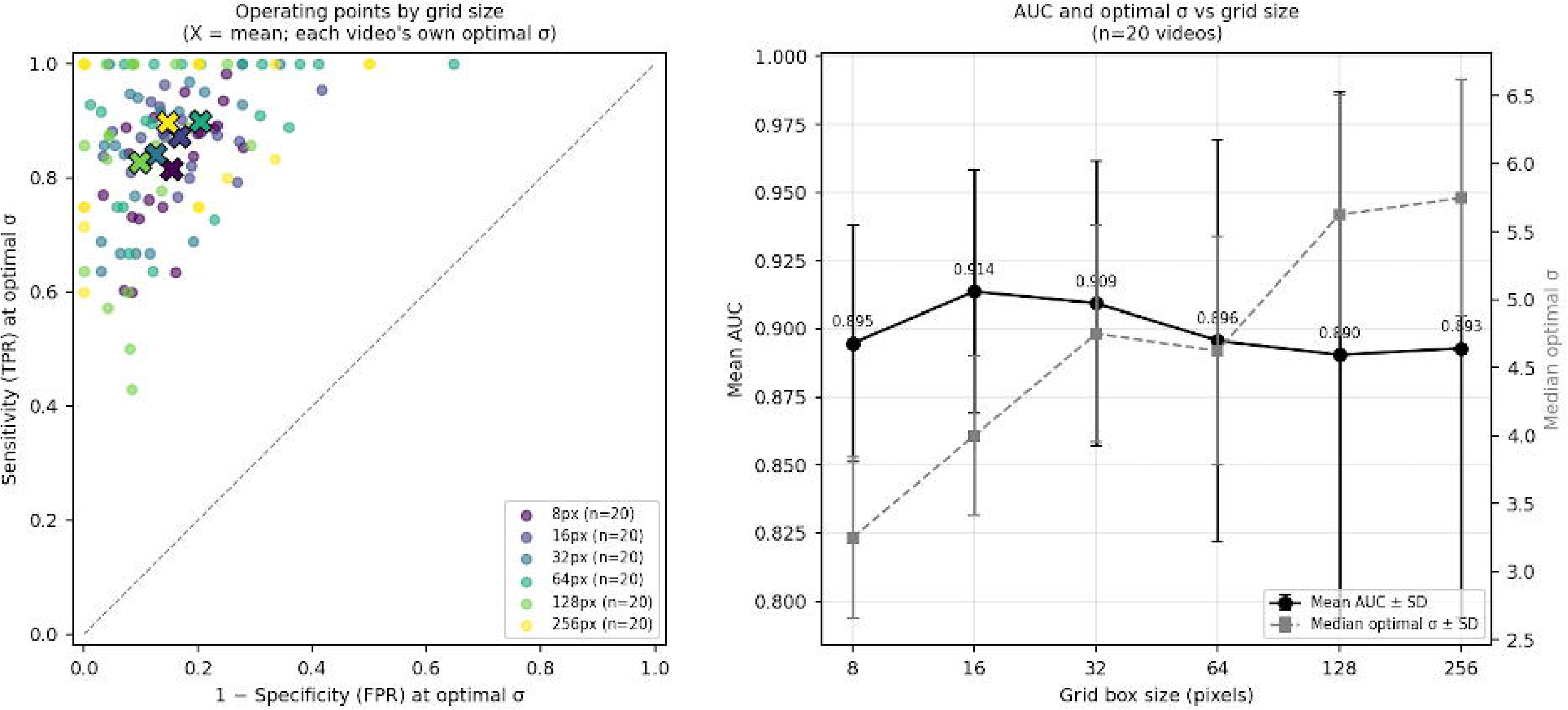
Grid-wise detection performance by box size. (A) Sensitivity and 1−specificity at each video’s individually optimal σ (Youden’s J), plotted separately for six grid box sizes (8– 256 px). Because only the optimal operating point was retained for each video rather than the full threshold sweep, discrete points rather than continuous ROC curves are shown; × markers denote the mean operating point per box size. (B) Mean AUC ± SD (black, left axis) and median optimal σ ± SD (gray, right axis) as a function of grid box size, aggregated across all 20 videos.

**Figure 6:**
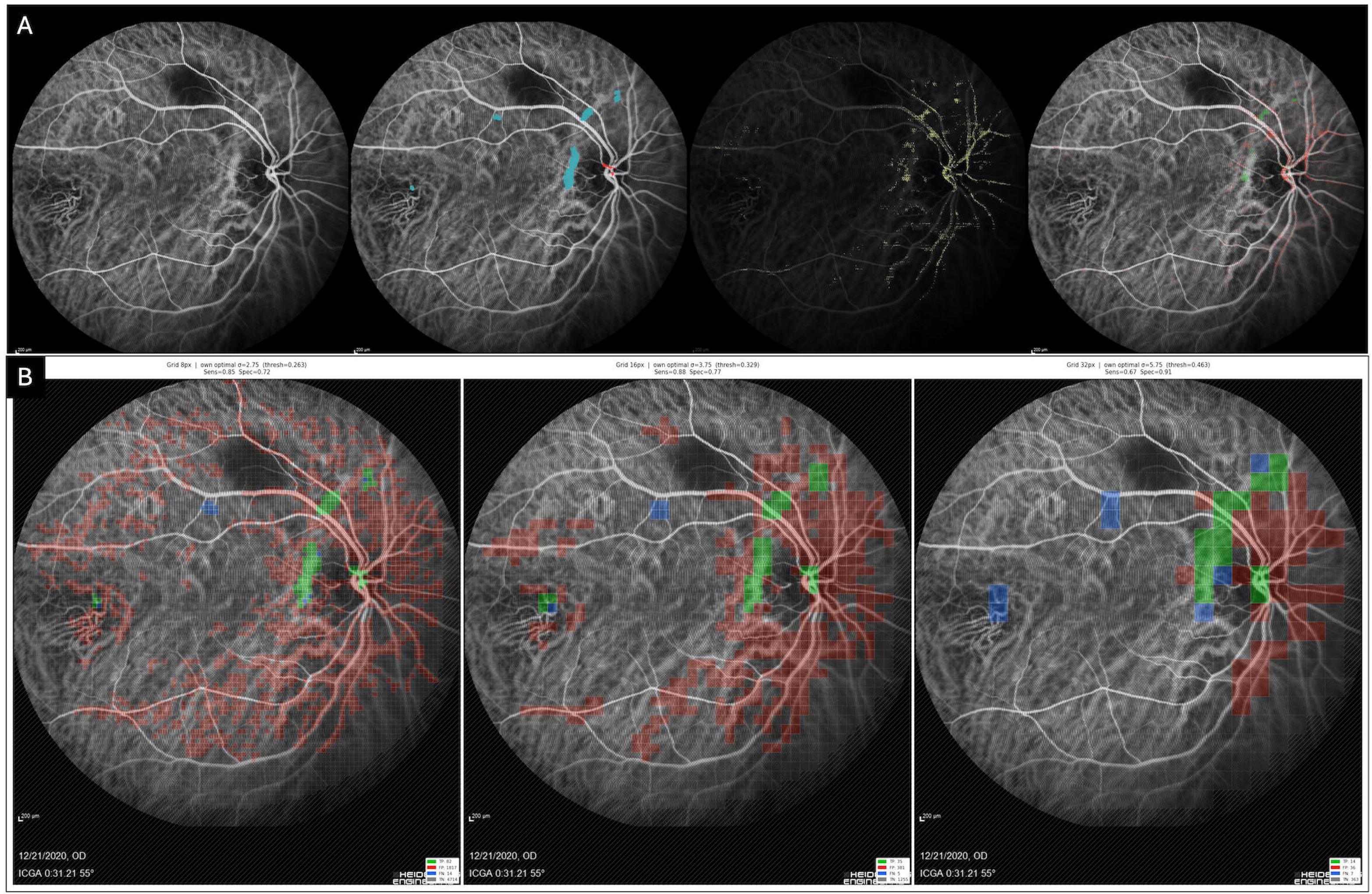
Detection accuracy for a representative video (Supplementary video 1). (A) Pixel-level matching (not used for analysis), evaluated at σ = 3.75. Panel 1: reference ICGA frame. Panel 2: grader consensus ground truth (primary, ≥2 of 4 annotation passes) (retinal pulsatile foci in red, choroidal pulsatile foci in cyan). Panel 3: algorithm-detected pulsatile pixels at this threshold, shown in yellow. Panel 4: pixel-wise agreement between panels 2 and 3. (B) Grid-level matching, evaluated separately at each box size’s own optimal σ (8px: σ=2.75; 16px: σ=3.75; 32px: σ=5.75 for this eye). Ground truth is the combined (retinal + choroidal) primary consensus. [Box/pixel color: green = true positive, red = false positive, blue = false negative, gray = true negative].

**Table 2.** Grid-wise receiver operating characteristics (ROC) (≥2/4 consensus, combined retinal+choroidal).

| Grid size | Median $\sigma$ | Mean $\sigma \pm$ SD | Mean AUC ± SD | Mean Sensitivity ± SD | Mean Specificity ± SD |
| --- | --- | --- | --- | --- | --- |
| 8px | 3.25 | $3.45 \pm 0.59$ | $0.895 \pm 0.043$ | $0.816 \pm 0.113$ | $0.848 \pm 0.073$ |
| 16px | 4.00 | $4.04 \pm 0.59$ | $0.914 \pm 0.045$ | $0.873 \pm 0.061$ | $0.832 \pm 0.090$ |
| 32px | 4.75 | $4.86 \pm 0.80$ | $0.909 \pm 0.052$ | $0.841 \pm 0.128$ | $0.876 \pm 0.087$ |
| 64px | 4.625 | $4.90 \pm 0.84$ | $0.896 \pm 0.074$ | $0.898 \pm 0.124$ | $0.796 \pm 0.161$ |
| 128px | 5.625 | $5.54 \pm 0.88$ | $0.890 \pm 0.097$ | $0.828 \pm 0.184$ | $0.903 \pm 0.076$ |
| 256px | 5.75 | $5.49 \pm 0.87$ | $0.893 \pm 0.099$ | $0.897 \pm 0.136$ | $0.854 \pm 0.172$ |

Area under the curve (AUC) by trapezoidal rule, mean ± SD across videos.
Sensitivity/specificity at Youden optimal

### Convergence and Final σ Recommendation

The detection-based recommendation (median of 16px and 32px grid-ROC optima) was σ = 4.00–4.75 (midpoint 4.38). The signal-derived recommendation (90% cumulative area reduction) was σ = 3.75 (Table 3, Figure 7). These two independently derived recommendations differed by 0.62σ, indicating strong agreement between a grader-based and an annotation-free validation approach. Thus, a σ value of 4.0, falling between the signal-derived (3.75) and grid-based (4.38) thresholds, could be used as the optimum σ for future studies.

**Figure 7.**
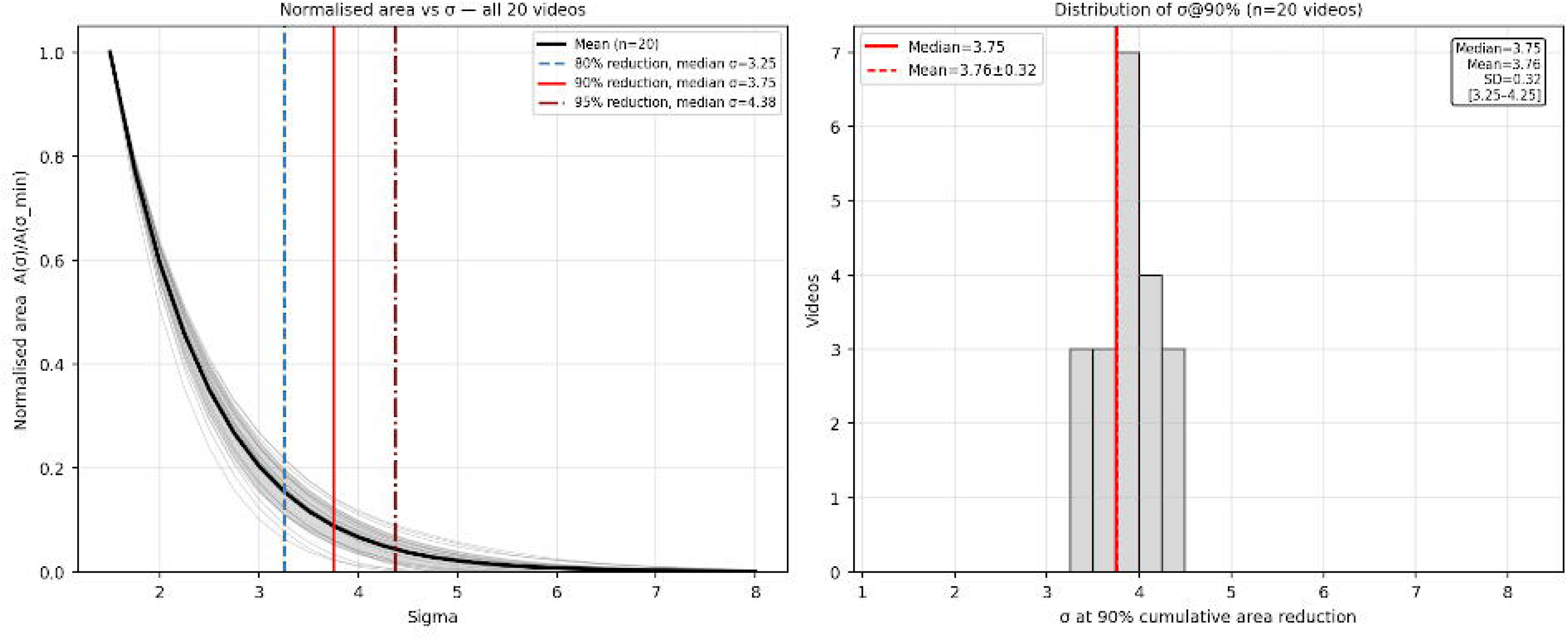
Area sensitivity analysis. Left: normalised area A(σ)/A(σ_min) for all 20 videos (grey) with mean ± SD (black); vertical lines at median σ for 80%, 90%, 95% reduction milestones. Right: distribution of σ@90% across 20 videos.

**Table 3.** Signal-derived sigma at each cumulative area reduction level.

| Reduction level | Median $\sigma$ | Mean $\sigma \pm$ SD |
| --- | --- | --- |
| 50% | 2.25 | $2.29 \pm 0.09$ |
| 70% | 2.75 | $2.77 \pm 0.18$ |
| 80% | 3.25 | $3.17 \pm 0.22$ |
| 90% (primary) | 3.75 | $3.76 \pm 0.32$ |
| 95% | 4.38 | $4.36 \pm 0.50$ |

## DISCUSSION

In this study, we developed and validated a computational algorithm for the automated detection and quantification of focal pulsation in video ICGA and demonstrated good performance compared with expert annotations. At a 16-pixel grid size, the pipeline achieved an AUC of 0.89, a sensitivity of 0.86, and a specificity of 0.80. Two independent validation methods converged on the same recommended detection threshold. Grader-based grid ROC analysis identified a median optimal sigma of 4.38 at grid level, and signal-based cumulative area reduction analysis independently identified sigma = 3.75 at the 90% reduction milestone. This converged at sigma of approximately 4.0. This sigma value indicates that the threshold that best separates grader-annotated pulsation regions from background noise also corresponds to the point at which most background signal has been eliminated. The pipeline simultaneously characterized retinal and choroidal pulsation foci, recorded their extent and amplitude, and output all findings as quantitative data, making them ready for integration into longitudinal studies.

The digital subtraction angiography method utilized by Cheung et al., in which sequential frames were subtracted to isolate the dye front, has improved our knowledge regarding choroidal pulsation.9 The authors used this approach in monkeys to demonstrate that the choriocapillaris fills in a pulsatile, lobular pattern, and later applied it in patients with CSCR to document oscillatory flow and pulsatile filling in pachyvessels. 9, 10 However, digital subtraction ICGA is limited to the dye-bolus phase of the angiogram and yields qualitative observations rather than quantitative metrics. Importantly, the method detects a moving dye front in the initial 5 to 10 seconds of the angiogram and is therefore prone to artifacts from early-phase brightness/contrast fluctuations and motion that occur when the operator adjusts the video to optimum contrast. The current pipeline addressed each of these limitations.

Rather than relying entirely on the unstable initial dye front, it automatically detects a stable section of the video and processes this stable phase of the angiogram to detect pulsation. By computing a composite score that multiplies local spatial phase coherence by oscillation amplitude at the cardiac frequency, the pipeline identifies pixels whose brightness genuinely oscillates with the heartbeat and is consistent with neighboring pixels, thereby distinguishing true pulsation from random noise, motion residuals, or single-pixel artifacts.

From a technical standpoint, the threshold σ is set as a number of standard deviations above each video’s own average synchrony score. This means the system adapts to the signal-to-noise ratio of each recording. This helps prevent false detections in noisy videos and missed detections in quiet ones. The semi-automatic method for choosing the heart rate, in which the pipeline suggests up to three possible frequencies and the operator selects the most practical one, adds a human check that helps prevent mistaking breathing, lateral movements of the video, or random harmonic signals for the heart rate. This was done to prioritize accuracy over full automation. However, fully automated frequency detection may be tried in the future.

At the grader level, pixel-level inter-grader agreement was fair, reflecting the fundamental difficulty of detecting and precisely delineating a pulsatile segment on a noisy video background. However, agreement improved substantially with different grid sizes, reaching moderate values by 16–32 pixels and good-to-excellent agreement at coarser grids. This progressive improvement with box size showed that graders agree on where pulsation occurs but not on its exact pixel-level extent. This justifies designing the validation using grid-wise rather than pixel-wise ROC. It is worth noting that inter-grader κ continued to rise well beyond the 16–32px range, reaching 0.75–0.81 for choroidal annotations at 128–256px. This was an expected phenomenon, where the κ values increase as regions of interest enlarge and absorb boundary-level disagreement. The moderate agreement levels indicate the unreliability of human measurements in detecting pulsation, underscoring the need for an automated detection algorithm.

It was interesting to see that the σ threshold sweep yielded different high AUC values across videos, primarily because the threshold varies with pixel fluctuations in each video. Thus, although in an ideal scenario this should be judged based on individual video quality, we sought a single sigma value that could be used universally across all videos to save time while minimizing compromise in detection sensitivity and specificity. Two independent approaches were used to identify an appropriate σ operating threshold. The detection-based approach used the grid-wise ROC results and provided optimal σ of 4.00 at 16px and 4.75 at 32px (mean value of 4.38). Although the AUC was high, the high false positives (Figure 6) suggest that the algorithm detected far more pulsation points than the graders did, underscoring the algorithm’s importance in future studies. The signal-derived approach was independent of any grader annotation and was based on cumulative pulsatile-area reduction, measured directly from the detection across multiple σ values, and reached 90% of its decline at a median σ of 3.75. These two recommendations differed by only 0.62. Because the two approaches draw on entirely different information (grader annotation in one and raw signal decay in the other), the mid-point value σ=4.0 (also coinciding exactly with the 16px grid-ROC optimum) is therefore the recommended σ value for future quantitative studies using this pipeline.

The ability to detect retinal vascular pulsation could open new clinical avenues. One of them could be the ocular perfusion pressure assessment for diseases such as glaucoma, diabetic retinopathy, and hypertension. In glaucoma, elevated intraocular pressure alters the pressure gradient between arterial and venous pressures, potentially affecting retinal vascular pulsation.11 Microvascular remodeling and loss of autoregulation in diabetic and hypertensive retinopathy could also affect the perfusion gradient and indirectly affect the pulsation. In each of these conditions, the amplitude and spatial distribution of retinal pulsation foci, as quantified by this pipeline, may offer information complementary to structural metrics. One important application would be the detection of spontaneous venous pulsation at the optic disc, which is present in approximately 90% of healthy individuals. Its absence is used as a clinically important early sign of raised intracranial pressure, preceding the development of papilloedema in many cases.12 Currently, detection of spontaneous venous pulsation relies entirely on subjective clinical observation. Because the pipeline detects oscillations at any pixel in the fundus image, it can, in principle, objectively identify and quantify spontaneous venous pulsation, albeit requiring dye angiography.

The choroidal application of this pipeline is where the most immediate clinical relevance lies. Diseases such as the pachychoroid spectrum are characterized by choroidal vascular hyperpermeability, dilated Haller layer, delayed choroidal venous filling, and vortex vein asymmetry.13 Cheung et al. demonstrated pulsatile filling of pachyvessels in watershed zones in CSCR eyes, with oscillatory flow reversal during diastole in the most severely affected cases.9 However, the authors could not quantify it, compare it between eyes, or track it longitudinally. By measuring pulsatile area and amplitude at each detected focus, it can characterize the spatial distribution of choroidal pulsation, thus addressing some unanswered questions from the previous studies. Beyond CSCR, the algorithm’s scope could naturally extend to polypoidal choroidal vasculopathy (PCV), as pulsatile filling has been documented on dynamic ICGA,14 thereby providing prognostic information about lesion activity and treatment response that static ICGA cannot capture.

There are several limitations of the current study that need to be acknowledged. First, the validation comprised only 20 eyes, acquired on a single platform, reducing the generalizability of the recommended sigma threshold, sensitivity, and specificity. Second, the cardiac frequency identification step is semi-automatic. This introduces an operator-dependent step that can affect reproducibility. Third, the pulsation amplitude, recorded in pixel-intensity units, does not indicate whether it is due to vessel wall motion or dye filling. Finally, this study does not validate quantification accuracy (area and amplitude) by comparing against an independent physiological ground truth, which could be extremely challenging in practice. The sensitivity analysis provides internal evidence that the measurement is stable and consistent across the sigma sweep. However, whether the absolute area values correspond to true pulsation area or magnitude remains to be established in future studies with appropriate physiological endpoints. Also, due to the retrospective nature of the study, the pulsation frequencies could not be validated against the patient’s true pulse rate, nor could they be validated against another video recording of the same eye captured consecutively to ensure consistency of output (which is practically impossible). Nonetheless, the study’s uniqueness lies in the first automated quantification of vascular pulsation, which could open new avenues for vascular metrics in ocular diseases.

In summary, we have created and validated an automated pipeline to detect and measure pulsation foci in retinal and choroidal vessels using dynamic ICGA videos. This method identifies pixels that oscillate in brightness in sync with the heartbeat. Two separate validation approaches, one involving expert grader annotations and the other based solely on signal distribution, both indicated the same optimal operating threshold, strongly supporting the robustness of the detection. The agreement analysis among graders also showed that the pipeline functions at a spatial scale consistent with where human experts agree on the presence of pulsation, at the grid level. Thus, pulsatile hemodynamics could add a new dimension to vascular metrics, pointing to features that static imaging biomarkers miss. These findings could have clinical relevance for conditions such as glaucoma, diabetes, hypertension, and the pachychoroid spectrum. Future studies should validate these results on larger, multicenter datasets across different imaging platforms and explore their potential as a prognostic biomarker.

## Data Availability

All data produced in the present study are available upon reasonable request to the authors

## Abbreviations

CSCR: Central Serous Chorioretinopathy
ICGA: Indocyanine Green Angiography
OCT: Optical Coherence Tomography
SD: Standard Deviation
ECC: Enhanced Correlation Coefficient
px: pixels
fps: frames per second
s: seconds
Hz: hertz
Bpm: beats per minute
PLV: phase-locking value
AUC: area under the curve
µm: micrometers
mm²: square millimeters
κ: kappa (Cohen’s kappa)
σ (sigma): threshold multiplier

---

Supplementary video 1: An Indocyanine Green Angiography (ICGA) of an eye with chronic central serous chorioretinopathy (left frame) shows clear pulsatile motion of the choroidal vessels (white arrows), identified as pulsation by the automated algorithm (right frame).

Supplementary video 2: An Indocyanine Green Angiography (ICGA) of the right eye of a patient with chronic central serous chorioretinopathy (left frame) shows clear pulsatile motion of the choroidal vessels (white arrow), identified as pulsation by the automated algorithm (right frame), with the brightest signal.

